# A methodology to generate epidemic scenarios for emerging infectious diseases based on the use of key calendar events

**DOI:** 10.1101/2022.04.29.22274465

**Authors:** M. Adrian Acuña-Zegarra, Mario Santana-Cibrian, Carlos E. Rodriguez Hernandez-Vela, Ramsés H. Mena, Jorge X. Velasco-Hernández

**Affiliations:** Departamento de Matematicas, Universidad de Sonora, Sonora, 83000, Mexico; Escuela Nacional de Estudios Superiores, Unidad Juriquilla, UNAM, Queretaro, 76230, Mexico; Instituto de Investigaciones en Matematicas Aplicadas y en Sistemas UNAM, CDMX 01000, Mexico; Instituto de Matematicas UNAM, Queretaro, 76230, Mexico

## Abstract

This work presents a methodology to recreate the observed dynamics of emerging infectious diseases and to generate short-term forecasts for their evolution based on superspreading events occurring on key calendar dates. The method is illustrated by the COVID-19 pandemic dynamics in Mexico and Peru up to January 31, 2022. We also produce scenarios obtained through the estimation of a time-dependent contact rate, with the main assumption that the dynamic of the disease is determined by the mobility and social activity of the population during holidays and other important calendar dates. First, historical changes in the effective contact rate on predetermined dates are estimated. Then, this information is used to forecast scenarios under the assumption that the trends of the effective contact rate observed in the past will be similar on the same but future key calendar dates. All other conditions are assumed to remain constant in the time scale of the projections. One of the main features of the methodology is that it avoids the necessity of fixing values of the dynamic parameters for the whole prediction period. Results show that considering the key dates as reference information is useful to recreate the different outbreaks, slow or fast-growing, that an epidemic can present and, in most cases, make good short-term predictions.

## Introduction

“Emerging” infectious diseases can be defined as infections that have newly appeared in a population or have existed but are rapidly increasing in incidence or geographic range [1]. More recent examples are H1N1 influenza (2009), Chikungunya (2014), Zika (2015), and COVID-19 (2019 to the present), the latter being the cause, so far, of more than 5.8 million deaths around the world.

Mathematical modeling has been widely used to study the COVID-19 epidemic. The transmission dynamics has been modeled with many different methodologies, several of them centered on estimating the effective reproduction number *R*_*t*_ with some version of the well-known Kermack-McKendrick model [2–5]. During 2020 and 2021, much effort was centered on projecting the COVID-19 pandemic and evaluating the efficacy of the mitigation strategies adopted to contain it [6, 7]. Around the world, the implementation of these measures has varied in strength ranging from strict and mandatory governmental enforcement to a voluntary personal decision. Regardless of the particular version of the mitigation strategy followed, these strategies must be based on local factors that combine public health status, economic impact and political conditions [8].

The central phenomena of human behavior that we explore are superspreading events occurring in particular calendar dates associated with religious, commercial, or civic holidays specific to the country. During or after these events (depending on their length), the contact rate changes. We, therefore, use these events as change points. We argue that in Mexico and Peru (both middle-income countries with a very stressed economic activity due to the pandemic), these change points (key calendar dates) reflect on the contact rate more clearly than changes imposed by the government (non-pharmaceutical interventions or NPIs) [9, 10]. Using key calendar dates has another advantage from the perspective of forecasting: these dates are known in advance and therefore mobility can be anticipated (short vacations, family visits, buying sprees, etc).

Here, we present a methodology that uses the known history of the disease reflected in the contact rate as a baseline to recreate the observed disease dynamics and forecast short-term epidemic scenarios. In [9], the authors have used a similar idea but, in our case, to determine changes in the contact rate, we look at particular events occurring on dates related to school, civic or religious periods (vacations, civic holidays, commercial events) that are known in advance each year for a given country. With this historical information in hand, we forecast the short-term evolution of the COVID-19 pandemic. Our results are illustrated considering as examples some Mexican states (Mexico City, Queretaro, Quintana Roo, and Sonora) and some departments of Peru (Arequipa, Cusco, Lima, and Piura).

The COVID-19 epidemic impact on the regional and global economy influences decisions on how and when businesses, public centers, tourism, schools and universities can safely reopen [11]. For decision-makers it has been of the highest importance to count with plausible scenarios for the evolution of the SARS-CoV-2 pandemic in order to design effective mitigation strategies. This knowledge is even more pressing in countries that lack the full infrastructure to acquire a more precise or, perhaps we should say, a less uncertain idea of the behavior of the pandemic for days or, ideally, weeks into the future. Our methodology is an advance in such an alternative.

The manuscript is organized as follows: First, we present the formulation of our mathematical model and the methodology used for parameter estimation. Then, we show our results for some Mexican states and departments of Peru. Finally, we present our discussion.

## Methods

### Mathematical model

A compartmental model is used to describe the evolution of the COVID-19 pandemic. The model considers three classes of infected individuals: Asymptomatic (I), Symptomatic (Y) and Reported (T). Once reported, infected individuals are effectively isolated and no longer participants in the transmission process. The model allows Susceptible (S) individuals to be Vaccinated (V) with a vaccination rate *ψ*. It is assumed that the vaccine is not perfect which implies that vaccinated people can be infected. Likewise, it is assumed that vaccinated individuals become susceptible after a certain period. Vital dynamics are also included since this work models the whole history of the pandemic. Fig 1 shows the corresponding model diagram.

**Fig 1.**
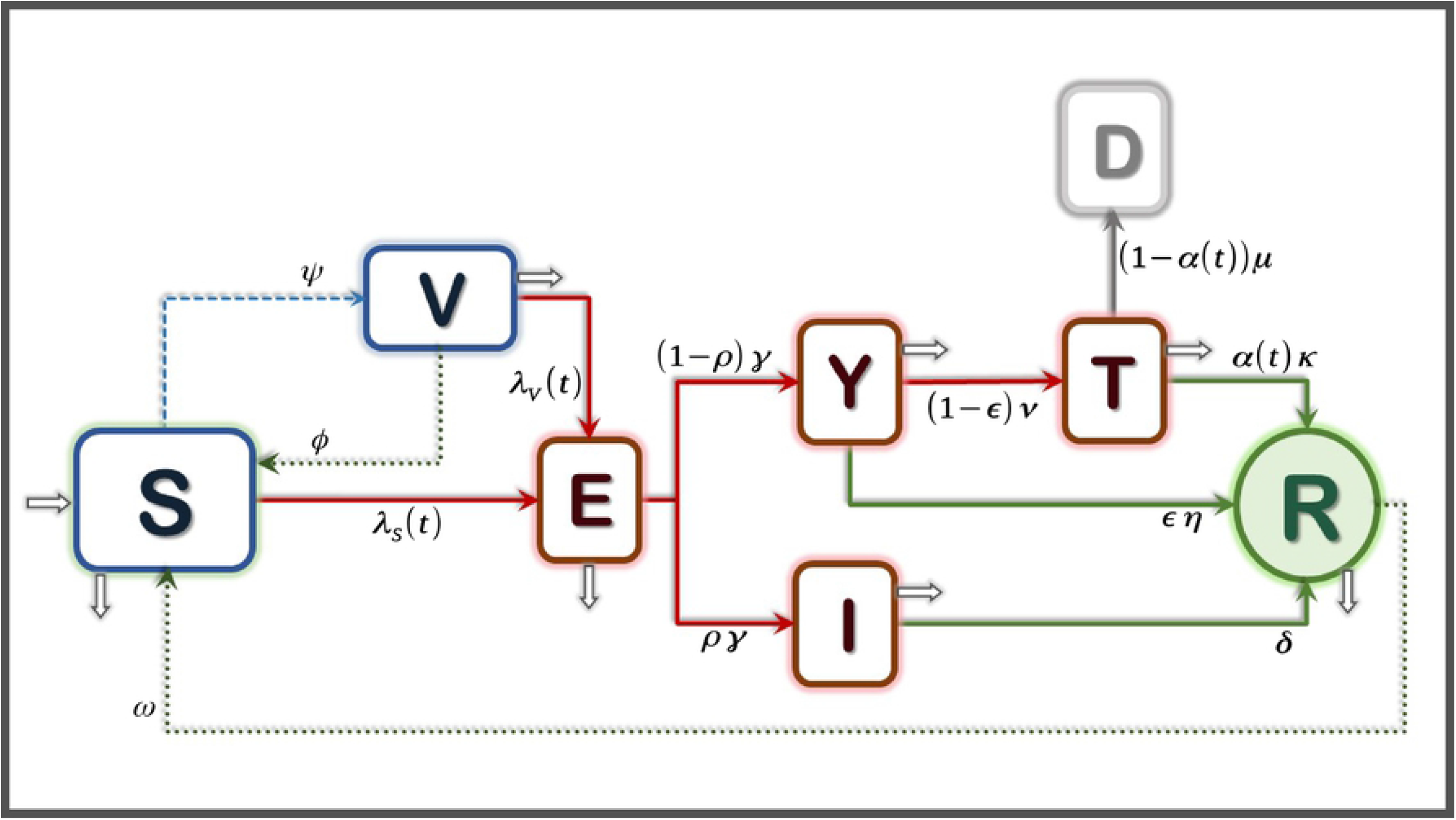
Mathematical model diagram. There are three types of infectious individuals: asymptomatic, symptomatic and reported. Reported cases do not play a role in transmission. Here, *λ*_*S*_(*t*) and *λ*_*V*_ (*t*) represent the infection force related to susceptible and vaccinated people, respectively.

Thus, the mathematical model is given by the following equations:

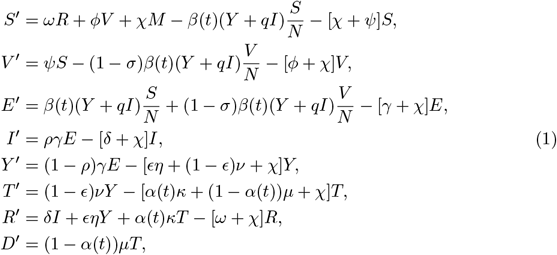

with *N* = *S* + *V* + *E* + *I* + *Y* + *R* and *M* = *N* + *T*. Table 1 shows a description of all the model’s parameters and their values.

**Table 1.** Description and values of the model’s parameters given in System 1. See [12, 13] for sources.

| Parameter | Description | Value | Units |
| --- | --- | --- | --- |
| $\psi$ | Vaccination rate | varying | days<sup>-1</sup> |
| $\nu$ | Screening rate | varying | days<sup>-1</sup> |
| $\mu$ | Disease mortality rate | varying | days<sup>-1</sup> |
| $\phi$ | Vaccine immunity waning rate | 0.006 | days<sup>-1</sup> |
| $\omega$ | Natural immunity waning rate | 0.006 | days<sup>-1</sup> |
| $\gamma$ | Incubation rate | 0.196 | days<sup>-1</sup> |
| $\delta$ | Asymptomatic recovery rate | 0.143 | days<sup>-1</sup> |
| $\eta$ | Symptomatic recovery rate | 0.071 | days<sup>-1</sup> |
| $\kappa$ | Reported recovery rate | 0.1 | days<sup>-1</sup> |
| $\chi$ | Natural mortality rate | $3.629 \times 10^{-5}$ | days<sup>-1</sup> |
| $\rho$ | Proportion of asymptomatic | 0.35 | % |
| $\epsilon$ | Proportion of symptomatic recovered | 0.94 | % |
| $q$ | Asymptomatic infectiousness reduction | 0.45 | % |
| $\sigma$ | Vaccine efficacy | varying | % |
| $\alpha(t)$ | Proportion of reported recovered people | estimated | % |
| $\beta(t)$ | Effective transmission contact rate | estimated | days<sup>-1</sup> |

The model depicted in Fig 1 is standard, but its main feature is how the contact rate *β* and the proportion of reported recovered people *α* are handled. Both parameters are time-dependent and defined through the interpolation of *k* change points. Interpolation is done using Hermite polynomials instead of splines to guarantee that both rates remain positive at any point in time. It is possible to estimate these two functions by using two time series: the number of reported cases and deaths. The *k* points in time when changes in *β* and *α* occur are pre-defined dates associated with the beginning of the mitigation measures and also civic or religious holidays.

#### Vaccination parameters

Many countries worldwide are using more than one vaccine. However, our model does not incorporate a detailed vaccination dynamic with different vaccines, doses, and efficacies since there is limited information about this process in Mexico.

The lack of information on the vaccination process affects the knowledge of important parameter values. Thus, as a first approach, it is assumed that susceptible individuals are vaccinated at a rate *ψ* to estimate the vaccination rate. Since the probability of having been vaccinated at time *t* is 1 *-* exp(*-ψt*) then, if a proportion *p* of the susceptible population is already vaccinated at time *T*_*V*_, then the vaccination rate that achieves this coverage is given by

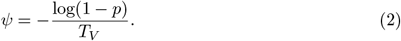

Parameters *p, T*_*V*_ and therefore, *ψ*, will vary between regions depending on the start of vaccination and vaccine stock. More details about the above will be provided in the Results Section.

### Parameter estimation

#### Data

We use the available COVID-19 data of each Mexican state [14]. Data consists of daily records of reported cases and deaths from the start of the pandemic (late February to early March, depending on the specific location) until February 7, 2022.

#### Statistical inference

A Bayesian approach is used to estimate key parameters of System 1: the time dependent contact rate *β*(*t*), the proportion of recovered reported individuals *α*(*t*) and the initial number of symptomatic and asymptomatic infected people, *I*_0_ and *Y*_0_, respectively.

To simplify the estimation process of functions *β*(*t*) and *α*(*t*), it is assumed that each of them is determined by their values at preset times *τ*_1_, *τ*_2_, …, *τ*_*k*_, which are associated to the key dates of the study regions. Let *a*_*i*_ be the proportion of reported recovered individuals at time *τ*_*i*_, and *b*_*i*_ the contact rate at time *τ*_*i*_, for *i* = 1, …, *k*. Values of *β*(*t*) and *α*(*t*) for any other point in time are obtained by interpolation using Hermite polynomials instead of splines to guarantee that both rates remain positive.

Let ***θ*** = (*E*_0_, *I*_0_, *Y*_0_, *a*_1_, *a*_2_, …, *a*_*k*_, *b*_1_, *b*_2_, …, *b*_*k*_) the vector of parameters that will be estimated. All the other parameters needed to solve System 1 are fixed and their values can be found in Table 1. It is assumed that initial conditions *E*_0_, *I*_0_ and *Y*_0_ can only take values in the interval (0,10). Parameters *b*_*i*_ are limited to the interval (0,5), while parameters *a*_1_ must take values in (0,1).

Let *W*_*j*_ and *X*_*j*_ be the random variables that count the number of daily COVID-19 reported infected individuals and deaths at time *t*_*j*_, respectively, for *j* = 1, 2, …*n*. Here, *t*_*j*_ represents the number of days since the start of the pandemic in each region. It is assumed that the probability distribution of *W*_*j*_, conditional on the vector of parameters ***θ***, is a Poisson distribution such that *E*[*W*_*j*_] = *C*(*t*_*j*_|***θ***) *- C*(*t*_*j-*1_|***θ***), with *C*(*t*|***θ***) being the cumulative number of reported cases according to System 1. Similarly, the probability distribution of *X*_*j*_, conditional on the vector of parameters ***θ***, is a Poisson distribution such that *E*[*X*_*j*_] = *D*(*t*_*j*_|***θ***) *- D*(*t*_*j-*1_|***θ***), with *D*(*t*|***θ***) being the cumulative number of deaths according to the compartimental model. Assuming that variables *W*_1_, *W*_2_, …, *W*_*n*_ and *X*_1_, *X*_2_, …, *X*_*n*_ are conditionally independent, then the likelihood function is given by

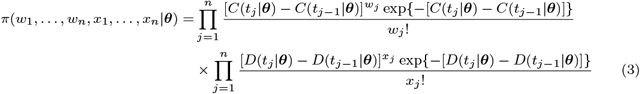

The joint prior distribution for vector ***θ*** is a product of independent Uniform distributions. For all the initial conditions *I*_0_ and *Y*_0_, the prior distribution is Uniform(0,10); for parameters *a*_1_, …, *a*_*k*_, the prior is Uniform(0,1); and finally, for *b*_1_, …, *b*_*k*_ the prior is Uniform(0,5). Then

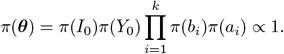

The posterior distribution of the parameters of interest is

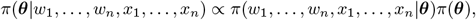

and it does not have an analytical form since the likelihood function depends on the numerical solution of the ODE System 1. We analyze the posterior distribution using an MCMC algorithm called *t-walk* [15] since it is implemented in the programming language Python and works well with highly correlated parameters, which typically appear in this type of estimation problems. For each state, 4 chains of 1,000,000 iterations are run, from which 200,000 are discarded as burn in. At the end, only 5000 iterations are retained to create the estimations presented in this work.

It is important to mention that confirmed COVID-19 cases can be grouped in different forms in order to represent the epidemic curve. In the case of Mexico, the number of cases can be grouped by date of symptoms onset, by the date when individuals seek medical attention (or tests), and by date of test results. In the case of Mexico, reported cases *X*_1_, …, *X*_*n*_ are those who seek medical attention at time *t*. This is why parameter *ν* is referred as the screening rate, the time from symptoms onset to testing. In the case of Peru, reported cases *X*_1_, …, *X*_*n*_ are those that got positive test results at time *t*. In that case, *ν*^*-*1^ represents the time from symptoms onset to test results. Notice that there is no need to modify the compartmental model to handle the difference between these two types of data, it is enough to change parameter *ν*.

## Results

### COVID-19 dynamics for selected Mexican states

As mention above, we use the available COVID-19 data of each Mexican state [14]. The dates when parameters *β* and *α* are assumed to change are shown in Table 2). These dates are the same for all states except for the start of the pandemic.

**Table 2.** Pre-defined dates used to estimate the contact rate and the proportion of reported recovered people for Mexican States.

| Description | Dates | Description | Dates |
| --- | --- | --- | --- |
| First case | Varying |  |  |
| NPIs start | 2020-03-23 | Valentines day | 2021-02-14 |
| Childrens day | 2020-04-30 | Easter | 2021-04-04 |
| Mothers day | 2020-05-10 | Childrens day | 2021-04-30 |
| Fathers day | 2020-06-21 | Mothers day | 2021-05-10 |
| Independence day | 2020-09-16 | Fathers day | 2021-06-20 |
| Day of the dead | 2020-11-02 | Independence day | 2021-09-16 |
| Buen Fin ends | 2020-11-21 | Day of the dead | 2021-11-02 |
| Day of the virgin | 2020-12-12 | Buen Fin ends | 2021-11-16 |
| Christmas | 2020-12-24 | Day of the virgin | 2021-12-12 |
| New Year | 2020-12-31 | Christmas | 2021-12-24 |
| Wise men day | 2021-01-06 | New Year | 2021-12-31 |
| Break | 2021-03-14 | Wise men day | 2022-01-06 |

To calculate the vaccination rate in Eq 2, let, for each state, *T*_*V*_ be the number of days from the start of vaccination up to February 7, 2022. The proportion of vaccinated people *p* during that period is approximated from official communications of the Mexican government [16]. On the other hand, we consider the total number of vaccines applied until February 7, 2022 [16], and the reported efficacy of the vaccines used in Mexico [17] to calculate the weighted vaccine efficacy (*σ*).

Fig 2 shows the COVID-19 model dynamics of reported cases in comparison with the observed data from the beginning of the pandemic to January 31, 2022. To exemplify the results, we chose states located at the north, center, and south of the country: Sonora, Mexico City, Queretaro, and Quintana Roo. Note that each epidemic curve has a different behaviour. Nevertheless, the proposed scheme based on key dates provides a good fit for the data.

**Fig 2.**
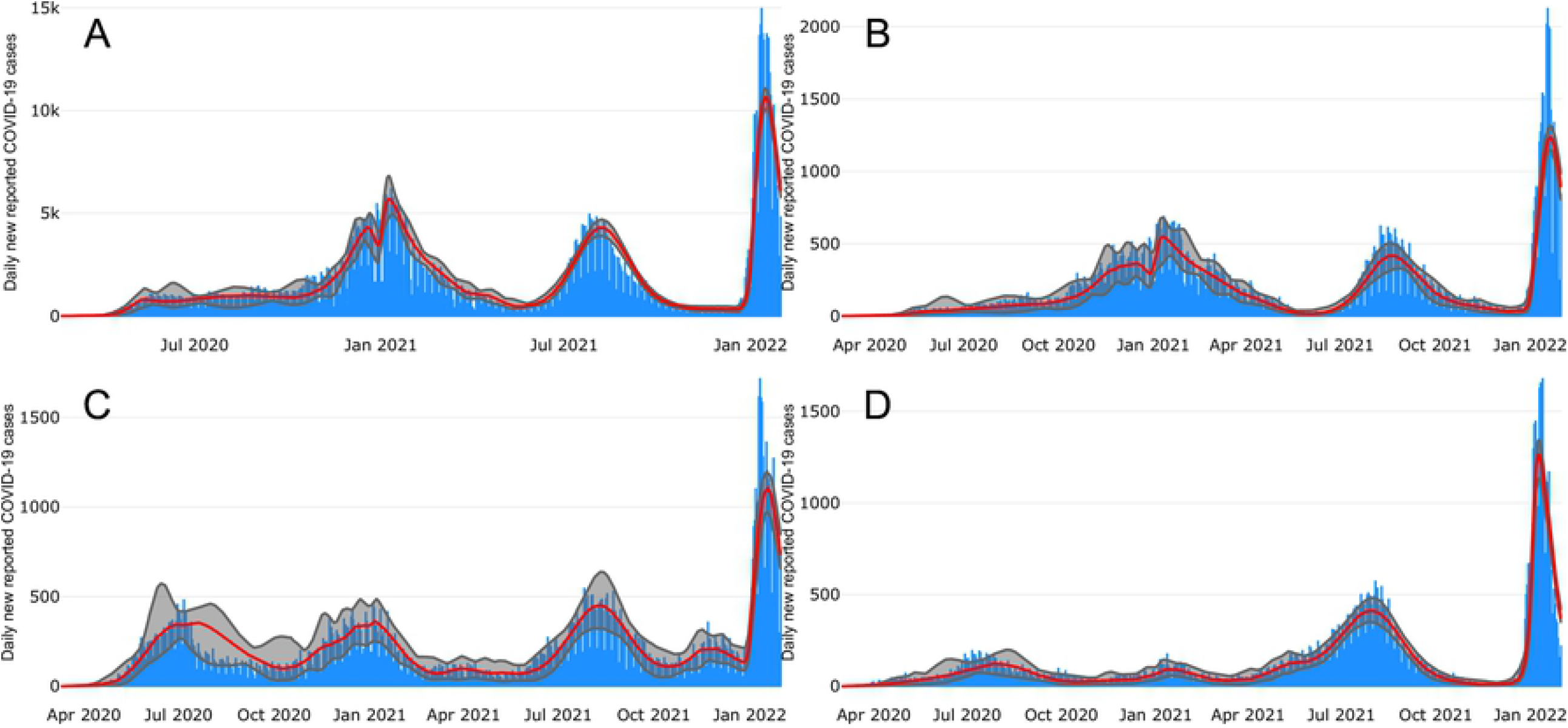
New reported COVID-19 cases in four Mexican states. (A) Mexico City, Queretaro, (C) Sonora, and (D) Quintana Roo. Blue bars show reported cases data. Red lines show the median posterior estimates. The gray shadow illustrates 95% pointwise probability regions.

Fig 3 shows the daily mortality given by the model in comparison with the observed deaths until January 31, 2022 for the same Mexican sates. The fit is also good even when the observed data shows state specific patterns. Furthermore, observe that the dynamics of reported cases and deaths is typical to each state.

**Fig 3.**
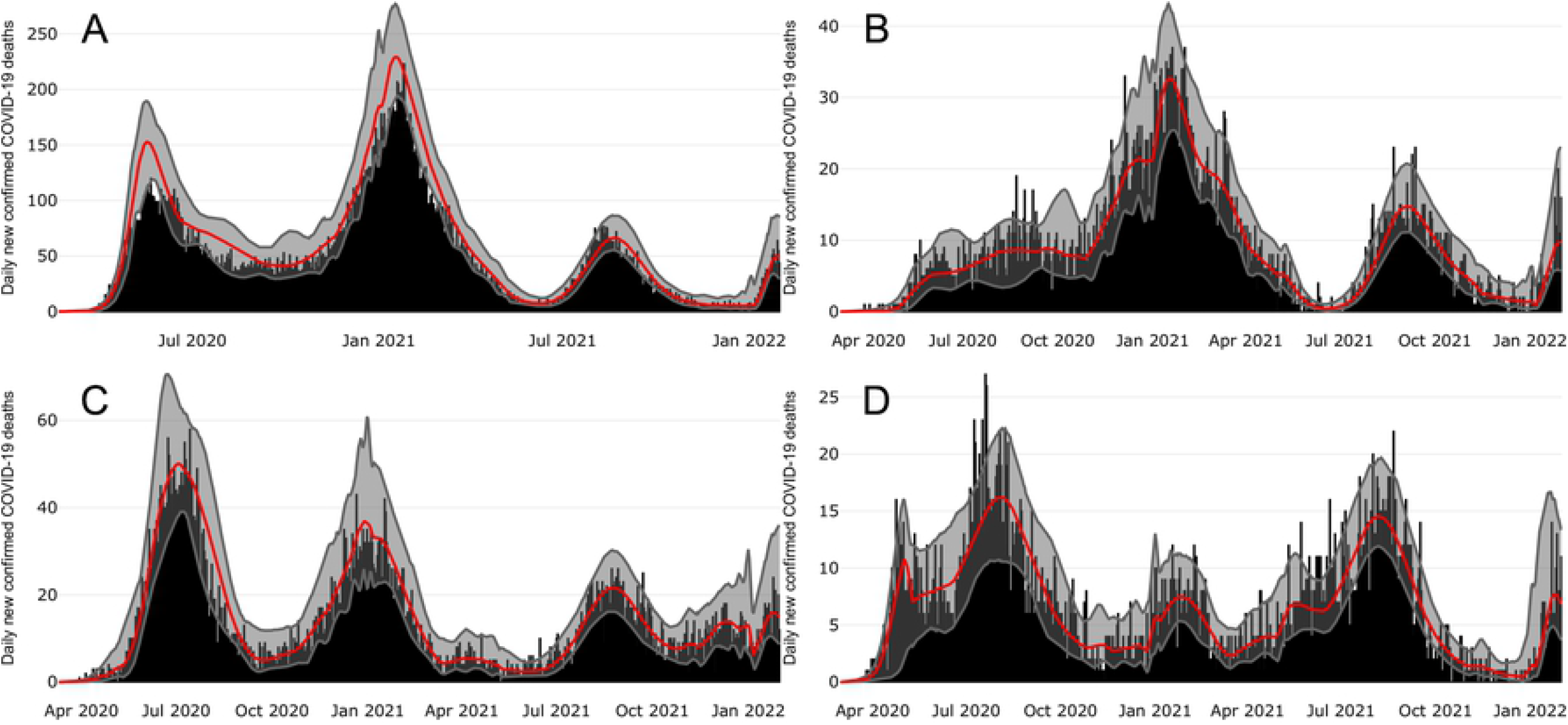
New reported COVID-19 deaths in four Mexican States. (A) Mexico City, (B) Queretaro, (C) Sonora, and (D) Quintana Roo. Black bars show reported deaths. Red lines show median posterior estimates. The gray shadow illustrates 95% pointwise probability regions.

### COVID-19 dynamics for Peru

We show here that the proposed methodology can be applied to other countries using COVID-19 data from Peru until February 7, 2022, as example [18, 19]. The key dates used to estimate parameters *β* and *α* for Peru are shown in Table 3. Data used to calculate values for *ψ* and *σ* can be found in [20].

**Table 3.** Pre-defined dates used to estimate the contact rate and the proportion of reported recovered people for department of Peru.

| Description | Dates | Description | Dates |
| --- | --- | --- | --- |
| First case | Varying |  |  |
| NPIs start | 2020-03-16 | Valentines day | 2021-02-14 |
| Easter | 2020-04-12 | Easter | 2021-04-04 |
| Labor day | 2020-05-01 | Labor day | 2021-05-01 |
| Mothers day | 2020-05-10 | Mothers day | 2021-05-10 |
| Fathers day | 2020-06-21 | Fathers day | 2021-06-20 |
| St Peter and St Paul day | 2020-06-29 | St Peter and St Paul day | 2021-06-29 |
| Independence day | 2020-07-28 | Independence day | 2021-07-28 |
| Santa Rosa de Lima day | 2020-08-30 | Santa Rosa de Lima day | 2021-08-30 |
| Battle of Angamos | 2020-10-08 | Battle of Angamos | 2021-10-08 |
| Saints' day | 2020-11-01 | Saints' day | 2021-11-01 |
| Immaculate Conception day | 2020-12-08 | Immaculate Conception day | 2021-12-08 |
| Christmas | 2020-12-24 | Christmas | 2021-12-24 |
| New Year | 2021-01-01 | New Year | 2022-01-01 |
| Wise men day | 2021-01-06 | Wise men day | 2022-01-06 |

Fig 4 and Fig 5 show observed and fitted COVID-19 confirmed cases and mortality, respectively, in four departments of Peru: Arequipa, Cusco, Lima and Piura. We can see that despite the differences between both countries, the fit continues to be good for confirmed cases and deaths.

**Fig 4.**
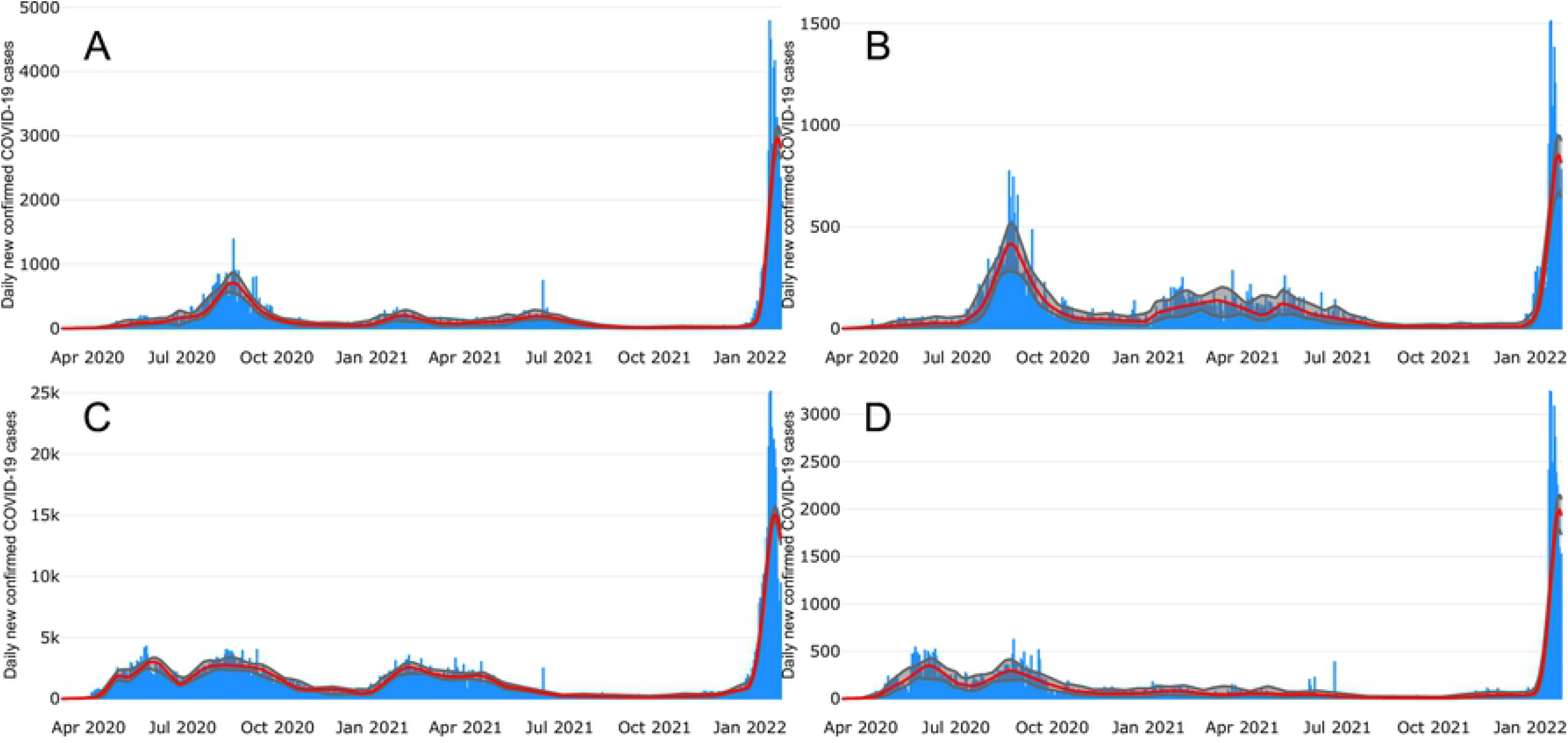
New confirmed COVID-19 cases in four departments of Peru. (A) Arequipa, (B) Cusco, (C) Lima, and (D) Piura. Blue bars shows confirmed cases. Red lines show the median posterior estimates. The gray shadow illustrates 95% pointwise probability regions.

**Fig 5.**
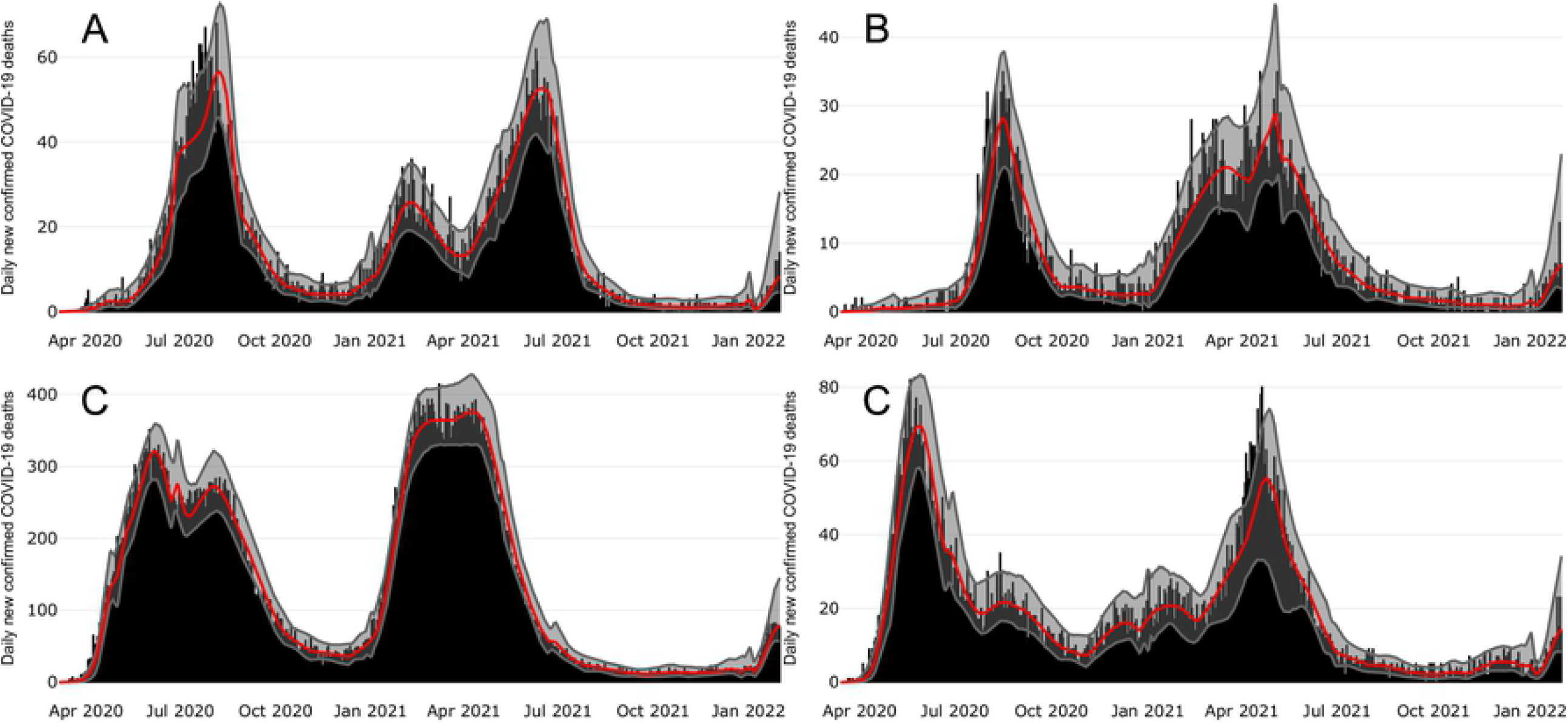
New reported COVID-19 deaths in some departments of Peru. (A) Arequipa, (B) Cusco, (C) Lima, and (D) Piura. Black bars shows reported cases data. Red lines show the median posterior estimates. The gray shadow illustrates 95% pointwise probability regions.

### Projected scenarios

Having *β* and *α* as time-dependent functions generates a good fit of the observed data. However, an important question is: how to predict the disease dynamics in the short, medium, and long term? In the case of models with dynamic parameters, it is necessary to set their values for the whole prediction period. The easiest way to address this is by continuing the last trend estimated from the observed data, which is what most models do. This can provide good predictions as long as there are no important events that impact transmission dynamics such as superspreading events, new virus variants, vaccination changes, climate changes, etc. This work proposes an alternative procedure to create predictive scenarios by using the history of the epidemic. The contact rate for the prediction period will change as it did during the same period of last year. For example, if the contact rate increased a 10% from December 24 to December 31, 2021, then it is assumed that it also will increase 10% from December 24 to December 31 during 2022. Finally, System 1 is solved using the new values of *β* to generate the the predicted state variables.

Fig 6 shows one month projections for Quintana Roo state at six different periods: June, July, August, September, October and November of 2021. Red bands show forecasts where darker tones denote more likely scenarios. Although there is a considerable amount of variability in some predictions, the observed trend is consistent with the forecast. Results for other states are shown in the Appendix A.

**Fig 6.**
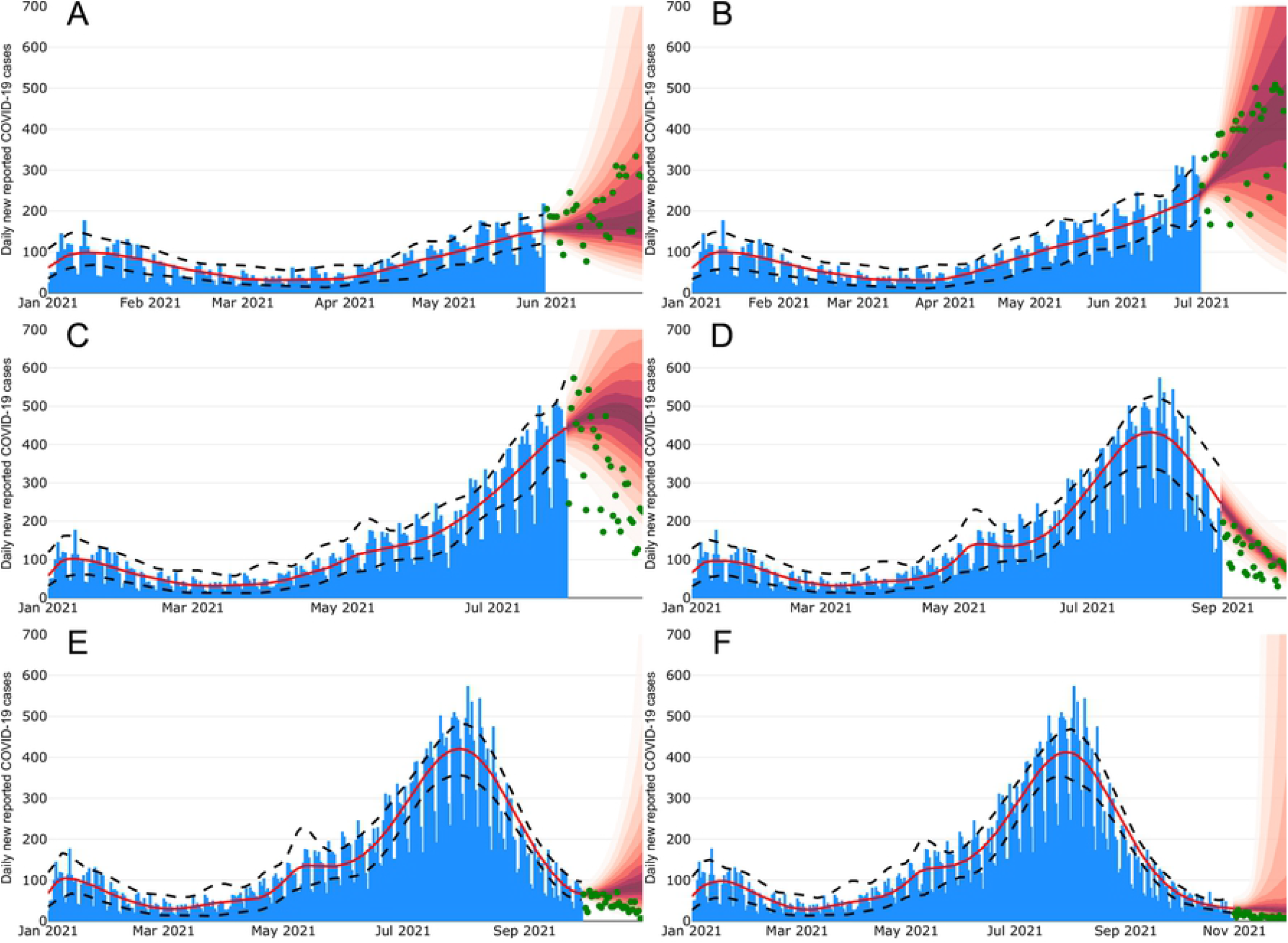
Projections of the COVID-19 evolution for Quintana Roo state. Each figure shows forecasts for different months: (A) June, (B) July, (C) August, (D) September, (E) October, (F) November. Blue bars show the reported data. Green dots represent observed data in the projection period. Red lines show the predictive posterior distribution for the dynamics of the reported cases. Darker tones indicate more likely scenarios.

## Discussion

Emerging infectious diseases are an important concern for public health. COVID-19 disease is the more recent example that has caused more than 5.8 million deaths and 419 million confirmed cases around the world as of late February, 2022 [21]. This disease has once again shown the role of mobility in the spread of acute respiratory diseases [13]. This is consistent with research from [22] where superspreading events are found to be phenomena that has shown to be determinant for many infectious diseases [22].

The epidemic dynamics observed in all of Mexico have been driven by events associated with heightened mobility and increased social activity [23–31] during holidays and other important calendar key dates. In previous works, earlier in the pandemic, it has been reported that key calendar days are a very good support to changes in the contact rates. As an example, in Mexico City the first case occurred by the end of February 2020, and the first set of mitigation measures was applied on March 23, 2020. Later that year, there were superspreading events in Mexico City during Easter celebrations (April 6-12) and early May (April 30 to May 10) that shifted the day of maximum incidence to the end of May, and pushed the epidemic into a quasi-stationary state characterized by values of *R*_*t*_ ≈ 1 [12, 13]. This behavior, observed around the world, has been explored in [32–34]. In particular, [34] claim that this quasi-linear growth and the maintenance of the effective reproduction number around *R*_*t*_ ≈ 1 for sustained periods of time, involves critical changes in the structure of the underlying contact network of individuals. In the case of Mexico City, we have argued that these changes relate to superspreading events on key calendar dates (Easter holidays and children’s, Holly Cross’ and mother’s days).

In this study, we have extended the ideas presented in [12, 13]. We have shown that key calendar days are a good reference to identify times where contact rates have changed. This has helped us to recreate different outbreaks of the COVID-19 disease dynamics in Mexico and Peru and also to give short-term projections.

Although this methodology has proven useful for fitting epidemic data, forecasting is still a challenge when extreme events affect the dynamic of the disease such as the appearance of new variants. Nonetheless, this is a problem that all modeling approaches have to face. The ideas presented in this work are intended to provide other perspectives to create predictive scenarios. The forecasting method proposed here are naive in the sense that it relies on the assumption that the behaviour observed in the past will be repeated in the future. Although this is a strong hypothesis, it still provides good short-term predictions in many situations as was shown. Moreover, we have shown that the method can capture important characteristics of the epidemic curve such as seasonality and the impact of repeated events such as holidays. Also, this procedure can potentially increase its relevance as the emerging disease moves towards and endemic state. Once two or more years of data are collected, projections could be done by a weighted average of the behaviour of the disease along several years. This is still a work in progress.

## Data Availability

All the data used in this work are available in the public domain: https://www.gob.mx/salud/documentos/datos-abiertos-152127 https://www.datosabiertos.gob.pe/group/datos-abiertos-de-covid-19?sort_by=changed&f%5B0%5D=field_tags%3A913&f%5B1%5D=field_tags%3A489

https://www.datosabiertos.gob.pe/group/datos-abiertos-de-covid-19?sort_by=changed&f%5B0%5D=field_tags%3A913&f%5B1%5D=field_tags%3A489

## Supporting information

**Appendix A Projected scenarios for other regions**. Figs 7 to 9 show one month projections for Mexico City, Queretaro state and Sonora state, respectively. Here, we consider six different periods: June, July, August, September, October and November of 2021. Red bands show forecasts where darker tones denote more likely scenarios.

**Fig 7.**
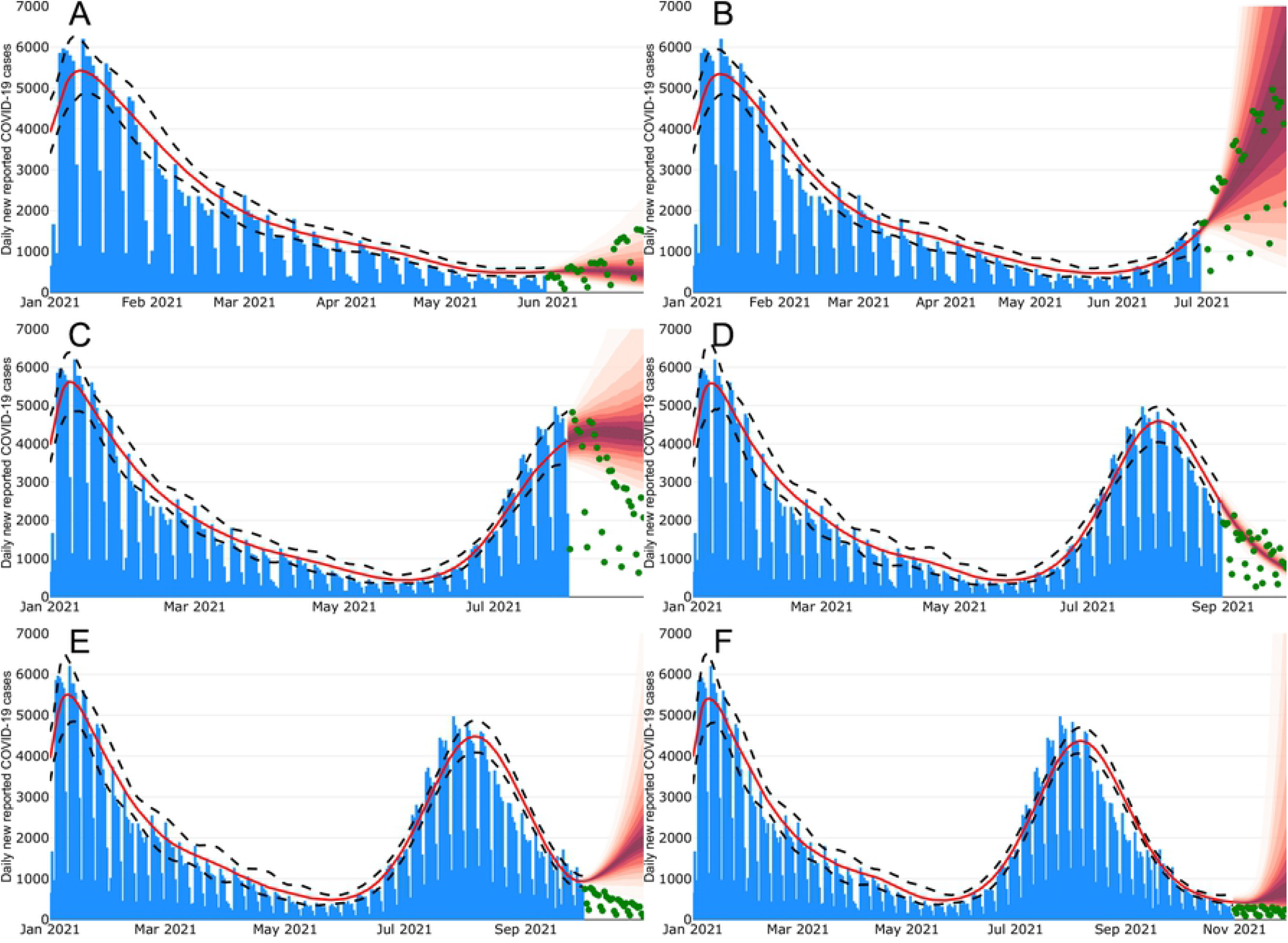
Projections of the COVID-19 evolution for Mexico City. Each figure shows the projections for different months. (A) June, (B) July, (C) August, (D) September, (E) October, (F) November. Blue bars show the reported data. Green dots represent data in the projection period. Red lines show the median posterior estimates for the dynamics of the reported cases. Red shadow, with different intensities, shows the projected scenarios.

**Fig 8.**
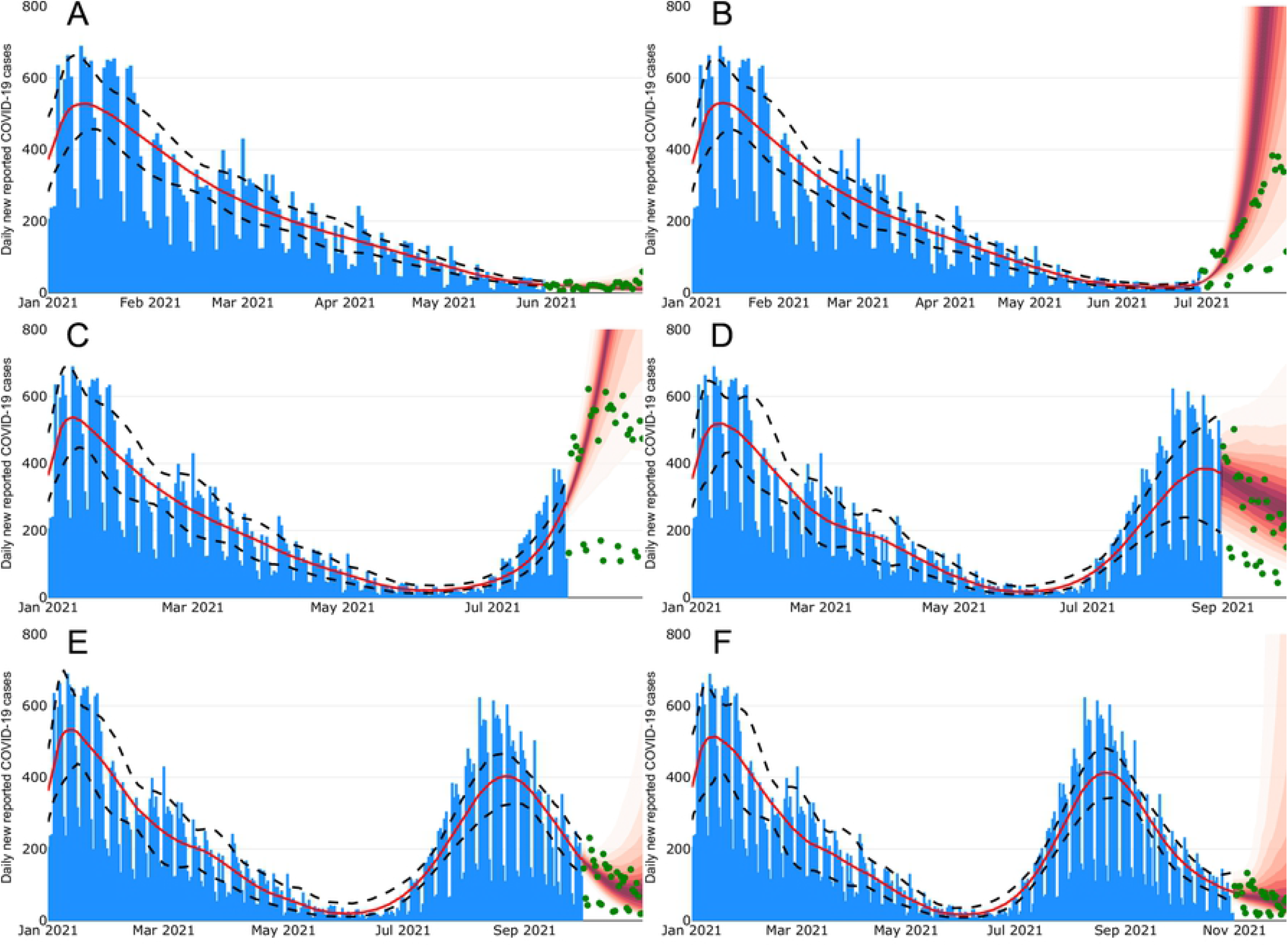
Projections of the COVID-19 evolution for Queretaro state. Each figure shows the projections for different months. (A) June, (B) July, (C) August, (D) September, (E) October, (F) November. Blue bars show the reported data. Green dots represent data in the projection period. Red lines show the median posterior estimates for the dynamics of the reported cases. Red shadow, with different intensities, shows the projected scenarios.

**Fig 9.**
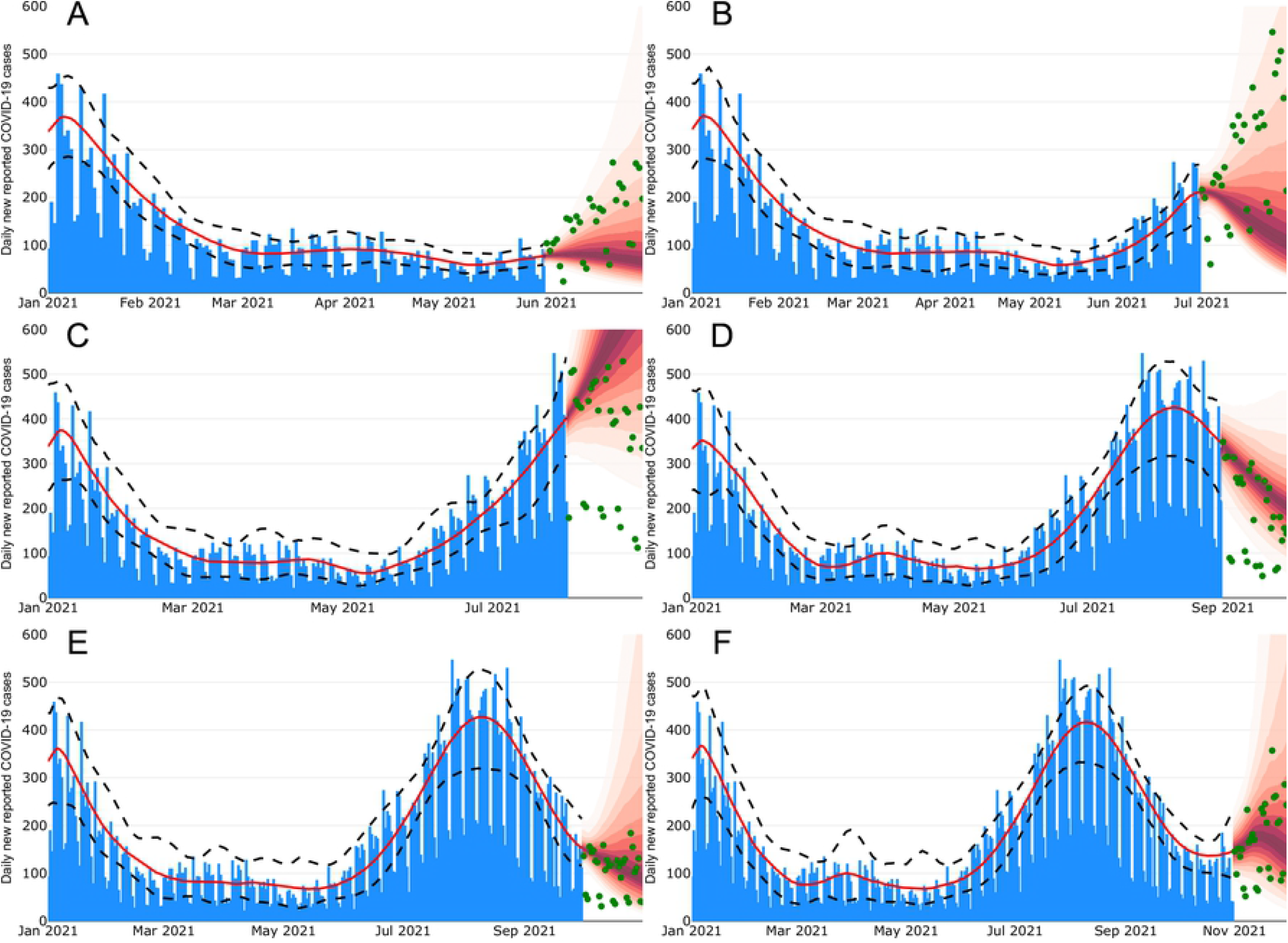
Projections of the COVID-19 evolution for Sonora state. Each figure shows the projections for different months. (A) June, (B) July, (C) August, (D) September, (E) October, (F) November. Blue bars show the reported data. Green dots represent data in the projection period. Red lines show the median posterior estimates for the dynamics of the reported cases. Red shadow, with different intensities, shows the projected scenarios.

## Acknowledgments

JXVH and MSC acknowledge support from UNAM PAPIIT DGAPA IN115720 and IV100220 grants. CERHV and RHM are grateful for the support of UNAM PAPIIT grant IG100221.

## Author Contributions

**Conceptualization:** Manuel A. Acuña-Zegarra, Mario Santana-Cibrian, Jorge X. Velasco-Hernández.

**Data curation:** Manuel A. Acuña-Zegarra, Mario Santana-Cibrian.

**Formal analysis:** Manuel A. Acuña-Zegarra, Mario Santana-Cibrian, Jorge X. Velasco-Hernández.

**Methodology:** Manuel A. Acuña-Zegarra, Mario Santana-Cibrian, Jorge X. Velasco-Hernández.

**Software:** Manuel A. Acuña-Zegarra, Mario Santana-Cibrian.

**Supervision:** Jorge X. Velasco-Hernández.

**Visualization:** Manuel A. Acuña-Zegarra, Mario Santana-Cibrian.

**Writing – Original Draft Preparation:** Manuel A. Acuña-Zegarra, Mario Santana-Cibrian, Carlos E. Rodriguez Hernandez-Vela, Ramsés H. Mena, Jorge X. Velasco-Hernández.

